# SARS-CoV-2 seroprevalence, vaccination, and hesitancy in agricultural workers in Guatemala

**DOI:** 10.1101/2022.02.22.22270907

**Authors:** Diva M. Calvimontes, Lyndsay Krisher, Alex Cruz-Aguilar, Daniel Pilloni-Alessio, Luis E. Crisostomo-Cal, Edgar A. Castañeda-Sosa, Jaime Butler-Dawson, Daniel Olson, Lee S. Newman, Edwin J. Asturias

## Abstract

**Background:** During the COVID-19 pandemic, serological tests to screen populations have provided better estimates of the cumulative incidence of infection. This study evaluated the seroprevalence of SARS-CoV-2 in agricultural workers in rural Guatemala, their COVID-19 vaccine uptake and vaccination attitudes.

**Methods:** A cross-sectional study was undertaken from August to November of 2021, in agricultural workers at a sugar plantation in Guatemala. A questionnaire was used to collect demographic, previous COVID-19 infection, vaccination, and attitudes toward vaccination. Serological testing was performed to detect SARS-CoV-2 IgM and IgG.

**Results:** Of the 4,343 study participants, 1,279 (29.4%) were seropositive for SARS-CoV-2 compared to 2.3% who reported previous COVID-19 infection. COVID-19 vaccine coverage was 85% for the first dose and 21.9% for second dose. Vaccine refusal was 0.6%, and 13.9% expressed some degree of vaccine hesitancy. Vaccine hesitant workers or those refusing were less likely to have had the COVID-19 vaccine. Main reasons to get the vaccine were to protect family, coworkers, and community.

**Conclusion:** Agricultural workers in countries like Guatemala have suffered a high incidence of asymptomatic and undetected SARS-CoV-2 infection. Most have received the COVID-19 vaccine, but there are moderate degrees of vaccine hesitancy that require better public health information to overcome it.

## Introduction

In January 2020, the World Health Organization (WHO) declared COVID-19 a public health emergency of international concern and by May 2020, WHO declared Latin America the epicenter of the COVID-19 pandemic. Latin America has seen some of the highest numbers of cases and deaths from COVID-19 in the world. As of December 31, 2021, more than 102 million confirmed cases of SARS-CoV-2 infection and 2.4 million have been reported in the region.^1^ However, these case counts underestimate the true cumulative incidence of infection. Serological tests to screen populations provide better estimates of the cumulative incidence of infection by complementing diagnostic tests for acute infection and helping to inform the public health response to COVID-19. A meta-analysis of seroprevalence studies carried out in various countries around the world showed that by 2020, seroprevalence was low in the general population (median 4.5%, IQR 2.4–8.4%).^2-4^ Most seroprevalence studies have been performed for convenience in urban populations. The objective of this study was to evaluate the seroprevalence of rural agricultural workers in Guatemala, the vaccination coverage against COVID-19, and the level of hesitancy and perception of these workers to vaccination.

## Methods

This cross-sectional epidemiological study was approved by the National Ethics Committee of the Ministry of Public Health and Social Assistance of Guatemala and considered a priority for public health. Agricultural workers over 18 years of age who applied to work at a sugar cane agro-industrial company from August to November 2021 were recruited. All participants signed an informed consent. Demographic data, underlying health, exposure to COVID-19 and severity of symptoms, dose of COVID-19 vaccine received, and perception of vaccination were collected through a standardized questionnaire. For the serological test, it was obtained from the residual blood sample of the routine selection tests carried out by the company. Serum samples were processed according to the manufacturer’s instructions for the STANDARD™ Q COVID-19 IgM/IgG Plus test (SD Biosensor, INC, South Korea). The STANDARD™ Q COVID-19 IgM/IgG Plus test is a rapid chromatographic immunoassay for the qualitative detection of specific antibodies (anti-SARS-CoV-2 nucleocapsid(N) protein antibodies and anti-SARS-CoV-2 Spike RBD antibodies) to SARS-CoV-2 and do not cross-react with the S-protein antibodies elicited by most vaccines. The reported sensitivity for IgM is 53.3% (36.1%; 69.8%) with a specificity of 100% (95.4%; 100%), for IgG the sensitivity is 73.3% (55.6%; 85.8%) and a specificity of 98.8%. (93.3%; 99.8%).^5-7^

### Data analysis

Participants were categorized as seropositive (IgM and/or IgG positive) or seronegative (IgM and IgG negative). Differences by age, sex, education, report of exposure to COVID-19 and vaccination were explored, estimating prevalence ratios (PR and 95% CI), and comparing means using t-test and categorical variables by chi-square (Stata 14.2, College Park, TX, USA).

## Results

A total of 4,498 workers were recruited, of which 4,343 (96.6%) consented to participate and a serological sample for SARS-CoV-2 antibodies was obtained. The demographic characteristics of the workers are shown in Table 1. The majority were men (99.5%), who worked in the sugar cane fields (99.9%), with an average age of 34 years, non-indigenous (99.7%), and from the surrounding areas where the company is located. The point prevalence of comorbidities was <1% with a low prevalence of smoking (2.4%). Ninety-seven (2.3%) of the workers reported having suffered from COVID-19 in the past, and of them 72 (74%) took a confirmatory diagnostic test (Nasopharyngeal PCR, n=35; antigen test, n=34). Compared to the 2.3% of self-reported previous COVID-19 infections, 1,279 (29.4%) workers were seropositive by IgG, IgM, or both. The probability that a worker seropositive for SARS-CoV-2 would report previous COVID-19 infection compared to those seronegative was 3.4 (OR 95% CI 2.3, 5.0; p<0.001). When asked about COVID-19 symptom severity, 17.8% reported not yet fully recovered from the disease (persistent fever, joint or muscle pain, anosmia, and persistent cough) and symptoms lasting between three to five weeks. Ninety-seven (2.3%) workers reported close relatives who had suffered from COVID-19; 40.2% of previously infected workers based on seropositive status also had a family member who had COVID-19 symptoms.

**Table 1.**
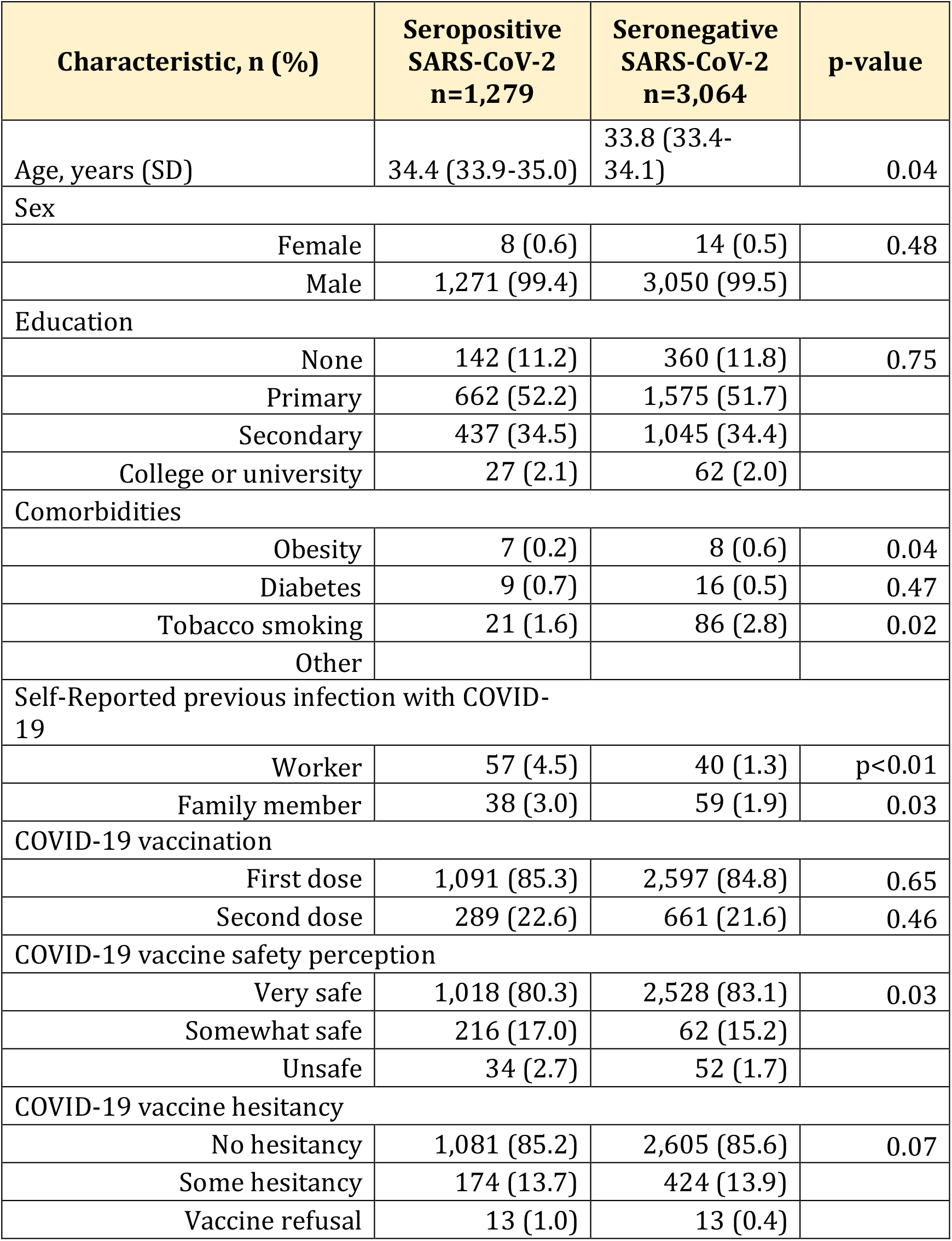
Characteristics of Agricultural Workers according to seropositivity for SARS-CoV-2 in Guatemala - 2021.

| Characteristic, n (%) | Seropositive SARS-CoV-2<br>n=1,279 | Seronegative SARS-CoV-2<br>n=3,064 | p-value |
| --- | --- | --- | --- |
| Age, years (SD) | 34.4 (33.9-35.0) | 33.8 (33.4-34.1) | 0.04 |
| Sex |  |  |  |
| Female | 8 (0.6) | 14 (0.5) | 0.48 |
| Male | 1,271 (99.4) | 3,050 (99.5) |  |
| Education |  |  |  |
| None | 142 (11.2) | 360 (11.8) | 0.75 |
| Primary | 662 (52.2) | 1,575 (51.7) |  |
| Secondary | 437 (34.5) | 1,045 (34.4) |  |
| College or university | 27 (2.1) | 62 (2.0) |  |
| Comorbidities |  |  |  |
| Obesity | 7 (0.2) | 8 (0.6) | 0.04 |
| Diabetes | 9 (0.7) | 16 (0.5) | 0.47 |
| Tobacco smoking | 21 (1.6) | 86 (2.8) | 0.02 |
| Other |  |  |  |
| Self-Reported previous infection with COVID-19 |  |  |  |
| Worker | 57 (4.5) | 40 (1.3) | p<0.01 |
| Family member | 38 (3.0) | 59 (1.9) | 0.03 |
| COVID-19 vaccination |  |  |  |
| First dose | 1,091 (85.3) | 2,597 (84.8) | 0.65 |
| Second dose | 289 (22.6) | 661 (21.6) | 0.46 |
| COVID-19 vaccine safety perception |  |  |  |
| Very safe | 1,018 (80.3) | 2,528 (83.1) | 0.03 |
| Somewhat safe | 216 (17.0) | 62 (15.2) |  |
| Unsafe | 34 (2.7) | 52 (1.7) |  |
| COVID-19 vaccine hesitancy |  |  |  |
| No hesitancy | 1,081 (85.2) | 2,605 (85.6) | 0.07 |
| Some hesitancy | 174 (13.7) | 424 (13.9) |  |
| Vaccine refusal | 13 (1.0) | 13 (0.4) |  |

Of the 4,343 workers, 85% had received the 1st dose of the COVID-19 vaccine at the time of the study (53% Moderna, 38% Sputnik-V, 9% AstraZeneca), and 21.9% the 2nd dose with the same vaccine antigens. No differences were observed in vaccination coverage according to SARS-CoV-2 seropositivity. A statistically significant but small proportion of SARS-CoV-2 seronegative workers expressed that the COVID-19 vaccine was very safe compared to seropositive workers (83.1 vs 80.3%; p=0.03).

The prevalence of vaccine refusal (not intending to get the vaccine) was 0.6%, and 13.9% of the workers showed some degree of vaccine hesitancy (stated by their intent to delay the decision to vaccinate, get vaccinated only as requirement, or having some insecurity about the vaccine). As expected, workers with no hesitancy were more likely to have the first or second dose of vaccine and over 20% of workers that were vaccine hesitant had received one or two doses. (Table 2) Among the workers with no hesitancy, protecting family, friends and co-workers were the most common reasons for getting the COVID-19 vaccine. Among workers who were hesitant, protecting family and friends and returning to work were the most common reasons, and among workers who refused 73% said they were unsure of a reason and 15% said to protect family and friends.

**Table 2.** COVID-19 vaccine uptake and reasons for intention to vaccinate according to level of hesitancy in agricultural workers in Guatemala, 2021.

| Characteristic, n (%) | No hesitancy | Some hesitancy | Vaccine refusal | p-value |
| --- | --- | --- | --- | --- |
| Number of participants | 3,686 (85.5) | 598 (13.9) | 26 (0.6) |  |
| **COVID-19 vaccine dose received** |  |  |  |  |
| First dose | 3,203 (86.9) | 483 (80.8) | 2 (7.7) | <0.001 |
| Second dose | 882 (24.2) | 58 (9.7) | 0 (0.0) | <0.001 |
| **Reason for getting the COVID19 vaccine** |  |  |  |  |
| Protect family and friends | 3,634 (98.6) | 312 (52.2) | 4 (15.4) | <0.001 |
| Protect co-workers | 2,154 (58.4) | 226 (37.8) | 2 (7.7) | <0.001 |
| Protect my community | 327 (8.9) | 63 (10.5) | 0 (0.0) | 0.12 |
| Return to work | 336 (9.1) | 306 (51.2) | 2 (7.7) | <0.001 |
| Return to social activities | 59 (1.6) | 26 (4.4) | 0 (0.0) | <0.001 |
| Travel | 6 (1.6) | 0 (0.0) | 0 (0.0) | 0.6 |
| Others induce me to get vaccinated | 47 (1.3) | 49 (8.2) | 2 (7.7) | <0.001 |
| Not sure | 6 (1.6) | 20 (3.3) | 19 (73.1) | <0.001 |
| **Sufficiency of COVID-19 vaccine information availability** |  |  |  |  |
| Yes | 2,111 (57.3) | 234 (39.1) | 4 (15.4) | <0.001 |
| Some | 1,427 (38.7) | 277 (46.3) | 18 (69.2) |  |
| No | 148 (4.0) | 87 (14.6) | 4 (15.4) |  |

Forty percent of workers reported that they had received some information about vaccines, and 241 (5.6%) that there was not enough information received. As shown in Table 2, most non-hesitant workers (57.3%) stated that there was enough information regarding the COVID-19 vaccines, compared to 39.1% of those with some hesitancy, and 15.4% of those refusing (p<0.001). Most workers trusted health care providers and the Ministry of Health to provide them with COVID-19 information, followed by their employer. In contrast, vaccine refusers had significantly more trust on their family or friends, or religious leaders to provide them with COVID-19 information. (Figure 1) The media (radio or television) were the least trusted source of vaccine information.

**Figure 1.**
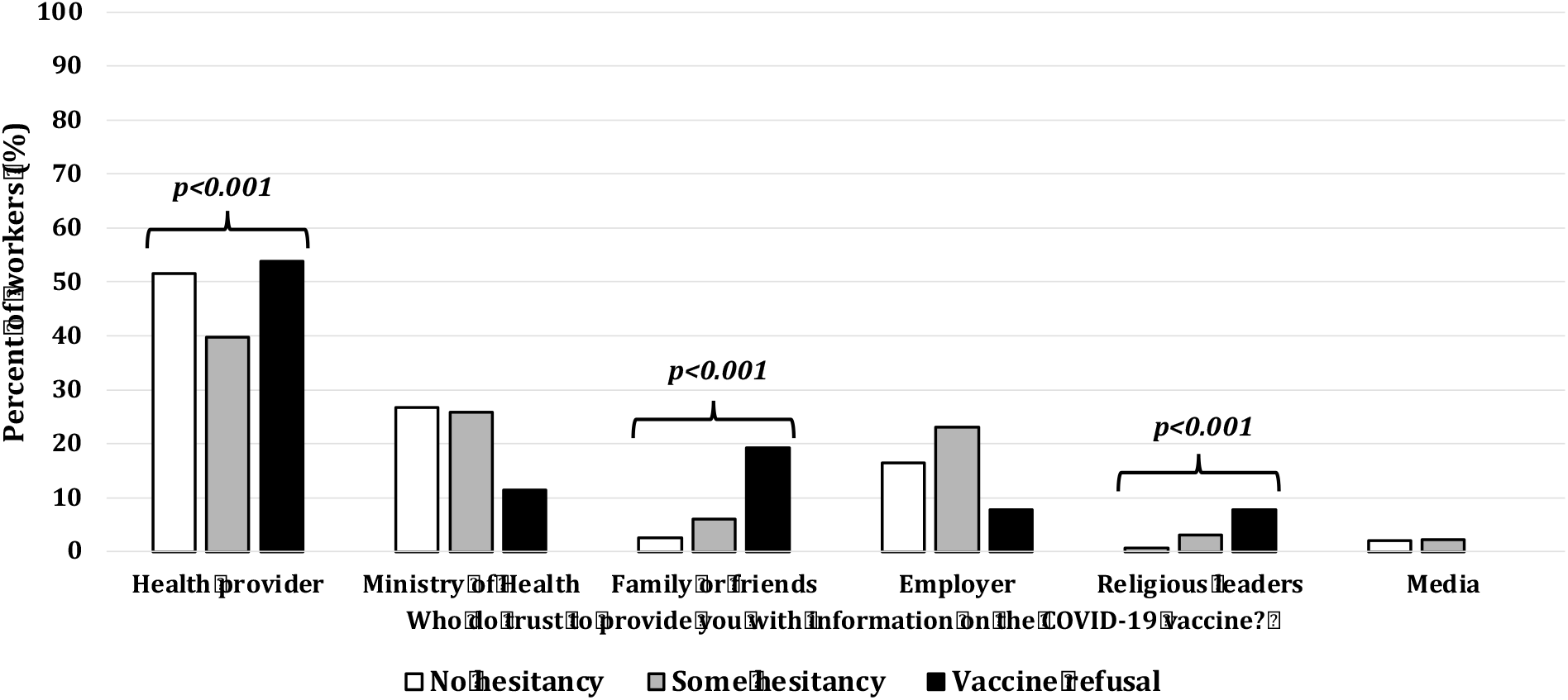
Trust of agricultural workers in COVID-19 information by source in rural Guatemala, 2021.

## Discussion

This is the first report of SARS-CoV-2 seroprevalence and COVID-19 vaccine decision-making in rural agricultural workers in a Latin American country. Guatemala, despite having achieved availability of COVID-19 vaccines, has managed by February 2022 to only vaccinate 41% of its population with 1 dose and 30% with 2 doses, with a large urban/rural disparity. Despite this, in agro-industrial companies, a high level of vaccination with the first dose and advances in the vaccination of the two-dose scheme are observed, probably due to the vaccination recommendations provided by employers to apply for work. Guatemala is among some countries in Latin America where vaccine mandates were not instituted.

This study shows that, in young populations of workers, there is a significant difference between the reporting of confirmed COVID-19 illness and SARS-CoV-2 seropositivity (2.3% vs 29.4%), possibly due to the frequency of asymptomatic illness in these populations and the limited availability of diagnostic tests for SARS-CoV-2. Even when three out of four workers who reported previous COVID-19 disease had taken some confirmatory test, most workers with mild upper respiratory infections, influenza like-illness (ILI), or exposed to a contact with COVID-19 infection do not get tested. In a recently published study by our team of a prospective cohort of banana farm workers in Guatemala showed that 25% of those who presented with ILI were positive for SARS-CoV-2 by PCR, and that the seropositivity for the year 2021 was 46%.^8^ Likewise, in several rural provinces in Peru, by March 2021, the seroprevalence was 59.0% (95% CI, 55 to 63%), confirming the intense spread of SARS-CoV-2 in areas where the lockdown and restrictions were less strict.^9^ The prevalence of COVID-19 vaccine refusal in this population was very low, but we were able to detect a moderate level of COVID-19 vaccine hesitancy similar to other studies in Latin America.^10^ These attitudes were directly linked to the probability of being vaccinated with 1 or 2 doses of COVID-19 vaccine and to the reasons and intentions for getting vaccinated. Our data also shows how COVID-19 vaccine information and communication impacted vaccine hesitancy, as most non-hesitant workers perceived that they had received enough vaccine information, especially from health care providers, the Ministry of Health, or their employer. In contrast, those who expressed some hesitancy or refused to get vaccinated, were more likely to express insufficient vaccine information, and to rely on their family, friends, or religious leaders who may have provided vaccine misinformation. Other studies in low and middle-income countries have shown that vaccine information is critical to dissipate fears and anxiety that have arose from the development of the COVID-19 vaccine and the misinformation spread thru the media and social platforms. ^11,12^

It is important to remark on some limitations of this study. First, the population surveyed does not represent other populations of agricultural and rural workers, and there is a selection bias towards young and healthy men, who were probably vaccinated given the recommendations provided by the employer before applying for a job. Second, this is a cross-sectional study, and most reports were based on self-reporting by workers which may have introduced recall bias. Despite this, the study provides valuable data for agricultural worker populations from middle-income countries that are considered essential for the economy and global food security.

## Data Availability

All data produced in the present study are available upon reasonable request to the authors

## Funding

The coordination of this study was funded by the Health and Education Policy Project Plus (HEP+) with funds from the United States Agency for International Development (USAID). The serological tests for SARS-CoV-2 (STANDARD Q COVID-19 IgM/IgG Plus Test) were donated by the Foundation for the Development of Guatemala – FUNDESA through the Ministry of Public Health of Guatemala.

## Acknowledgments

Our team would like to thank Pantaleon Foundation nurses and personnel who participated in the data and sample collection. We also will like to thank Juan Carlos Zapata at FUNDESA for his support and donations for this study, and Cesar Conde, MS for his technical advice and support at the National Laboratory of the Ministry of Public Health.

## Conflicts of interest

The listed authors certify that they have no affiliation with or involvement in any organization or entity with any financial interest, or non-financial interest in the subject matter or materials discussed in this manuscript.

## Notes

### Competing Interest Statement

The authors have declared no competing interest.

### Funding Statement

Health and Education Policy Project Plus (HEP+)
United States Agency for International Development (USAID)
Foundation for the Development of Guatemala (FUNDESA)

### Author Declarations

National Ethics Committee of Ministry of Public Health and Social Welfare of Guatemala gave ethical approval for this work

